# EpiKG2DAG: a Framework for Automated DAG Construction from Biomedical Text

**DOI:** 10.64898/2026.08.09.26360023

**Authors:** Guanghui Deng, Jian Du

**Affiliations:** Institute of Medical Technology, Peking University Health Science Center, Beijing, 100191, China; National Institute of Health Data Science, Peking University, Beijing, 100191, China

**Keywords:** Knowledge graph, Directed acyclic graph, Evidence Retrieval, Causal inference, Epidemiology

## Abstract

While Directed Acyclic Graphs (DAGs) are essential for causal inference, their construction often relies on expert heuristics, which bypasses systematic evidence synthesis and creates a critical “evidence retrieval gap” in causal modeling. This study introduces EpiKG2DAG, a framework that supports evidence-anchored candidate DAG generation by transforming unstructured biomedical abstracts into structured epidemiological associations. We utilized DeepSeek-V3 to extract exposure–outcome association triplets from 189,266 abstracts and employed SapBERT for semantic normalization against UMLS concepts. The resulting Epidemiological Knowledge Graph (EpiKG) enables the automated identification of candidate confounders, mediators, and colliders based on graph-theoretic motifs and literature-derived evidence. A case study on COVID-19 and AKI demonstrates that the framework uncovers non-obvious confounders, such as air pollution, while ensuring evidence traceability. This work contributes to the field by mitigating the knowledge-acquisition bottleneck and providing a transparent, reproducible foundation for evidence-based causal modeling.

## INTRODUCTION

Directed Acyclic Graphs (DAGs) are increasingly used to represent causal assumptions and guide confounder identification in epidemiological research (Feeney et al., 2025) However, constructing DAGs typically relies on researchers’ prior knowledge, manual literature consultation, and expert judgment, which becomes increasingly challenging as biomedical literature grows rapidly and relevant variables become more complex. In DAGs, three types of third-party variables are particularly important for a target exposure–outcome pair: common causes, intermediate variables, and common effects, corresponding to candidate confounders, mediators, and colliders, respectively. Manually building such a graph with distinct structural information is time-consuming, error-prone, and unsystematic for researchers(Bai et al., 2024; Malec et al., 2023). Consequently, it is imperative to extract epidemiological associations from empirical findings in scientific literature to build the evidence-based DAG.

The challenge of constructing reliable DAGs is rooted not only in the complexity of variable selection but also in a fundamental evidence retrieval gap. Recent meta-epidemiological evidence suggests a “credibility crisis” in causal modeling: approximately 70% of DAGs published in the four leading medical journals fail to report whether any systematic literature retrieval was conducted during their development(Deng & Du, 2026). This widespread lack of transparent evidence retrieval and documentation suggests that many causal structures may rely on subjective heuristics or fragmented knowledge rather than clearly traceable emporocal evidence. To address this gap, we frame candidate DAG construction as an evidence retrieval and knowledge organization task. Our framework retrieves findings from three study designs commonly used to generate or evaluate causal claims in epidemiolog: Randomized Controlled Trials (RCTs), Cohort Studies, and Mendelian Randomization (MR).These study designs provide a relevant evidence corpus for identifying candidate causal structures, while not implying that every extracted association represents definitive causal evidence.

In this study, we present EpiKG2DAG, an end-to-end framework designed to bridge the gap between large-scale literature retrieval and structural causal modeling. By leveraging Large Language Models (LLMs) for high-throughput information extraction and employing SapBERT for semantic normalization across standardized ontologies, the framework transforms 189,266 multi-source abstracts into a structured Epidemiological Knowledge Graph (EpiKG). Our approach facilitates the automated identification of candidate third-party variables—potential confounders, mediators, and colliders—based on literature-derived evidence. Through a case study on the causal relationship between COVID-19 and Acute Kidney Injury (AKI), we demonstrate that this framework can reduce the knowledge burden on researchers and uncover non-obvious confounders that may be overlooked in expert-driven models. Ultimately, EpiKG provides a transparent, reproducible, and evidence-anchored foundation for candidate DAG construction in the era of big data.

## CONCEPTUAL FOUNDATION OF EPIKG2DAG

The conceptual foundation of EpiKG2DAG is grounded in the graph-theoretic representation of causal assumptions using directed acyclic graphs (DAGs). A causal DAG can be formally defined as a graph (*G* = (*V,E*)), where (*V* = {*v*_1_, …, *v*_*n*_}) denotes a set of variables and (E) denotes a set of directed edges among these variables. Each directed edge (*v*_*i*_ → *v*_*j*_) represents an assumed causal influence from (*v*_*i*_) to (*v*_*j*_), and the graph is acyclic in the sense that no variable can be reached again by following a sequence of directed edges. Causal DAGs provide a formal language for representing assumptions about causal structure and have been widely used in epidemiology to identify variables that should or should not be adjusted for when estimating causal effects (Feeney et al., 2025; Greenland et al., 1999; Hernán et al., 2002).

In this study, we operationalize three DAG-based structural roles that are central to epidemiological causal reasoning: confounders, mediators, and colliders. For a given exposure (*X*) and outcome (*Y*), a candidate confounder is represented as a common cause (*C*) that has directed edges toward both (*X*) and (*Y*), that is, (*C* → *Y*) and (*C* → *Y*). A candidate mediator is represented as an intermediate variable (*M*) on a directed pathway from the exposure to the outcome, that is, (*X* → *M* → *Y*). A candidate collider is represented as a common effect (*K*) of the exposure and outcome, that is, (*X* → *K* ← *Y*). These three structures are important because they imply different analytical decisions: confounders are often considered for adjustment to reduce confounding bias, mediators may lie on the causal pathway of interest, and conditioning on colliders may introduce selection or collider-stratification bias (Greenland et al., 1999; Pearl, 2009; Textor et al., 2016).

EpiKG2DAG translates this graph-theoretic foundation into a computable evidence-retrieval framework. Literature-derived exposure–outcome associations are first organized as directed edges in the Epidemiological Knowledge Graph (EpiKG), with each edge linked to its source publications. Given a user-specified exposure–outcome pair, the framework searches the graph for candidate common causes, intermediate variables, and common effects according to their topological relationships with (*X*) and (*Y*). These graph motifs are then assembled into an evidence-anchored candidate DAG to support subsequent causal modeling.

## METHODS

The overall architecture of EpiKG2DAG is shown in Figure 1. The framework consists of three sequential modules: Evidence Retrieval Engine, Semantic Normalization and EpiKG Construction, and DAG Synthesis and Decision Support. First, the Evidence Retrieval Engine searches PubMed using predefined study-design and topic filters and then applies a large language model to extract structured exposure–outcome associations from titles and abstracts. Second, the Semantic Normalization and EpiKG Construction module maps heterogeneous biomedical terms to standardized UMLS concepts using SapBERT and organizes the extracted associations into the Epidemiological Knowledge Graph. Third, the DAG Synthesis and Decision Support module identifies candidate confounders, mediators, and colliders for a user-specified exposure – outcome pair according to the graph-theoretic definitions described above. Each edge remains linked to its source publications, enabling evidence tracing and expert verification.

**Figure 1.**
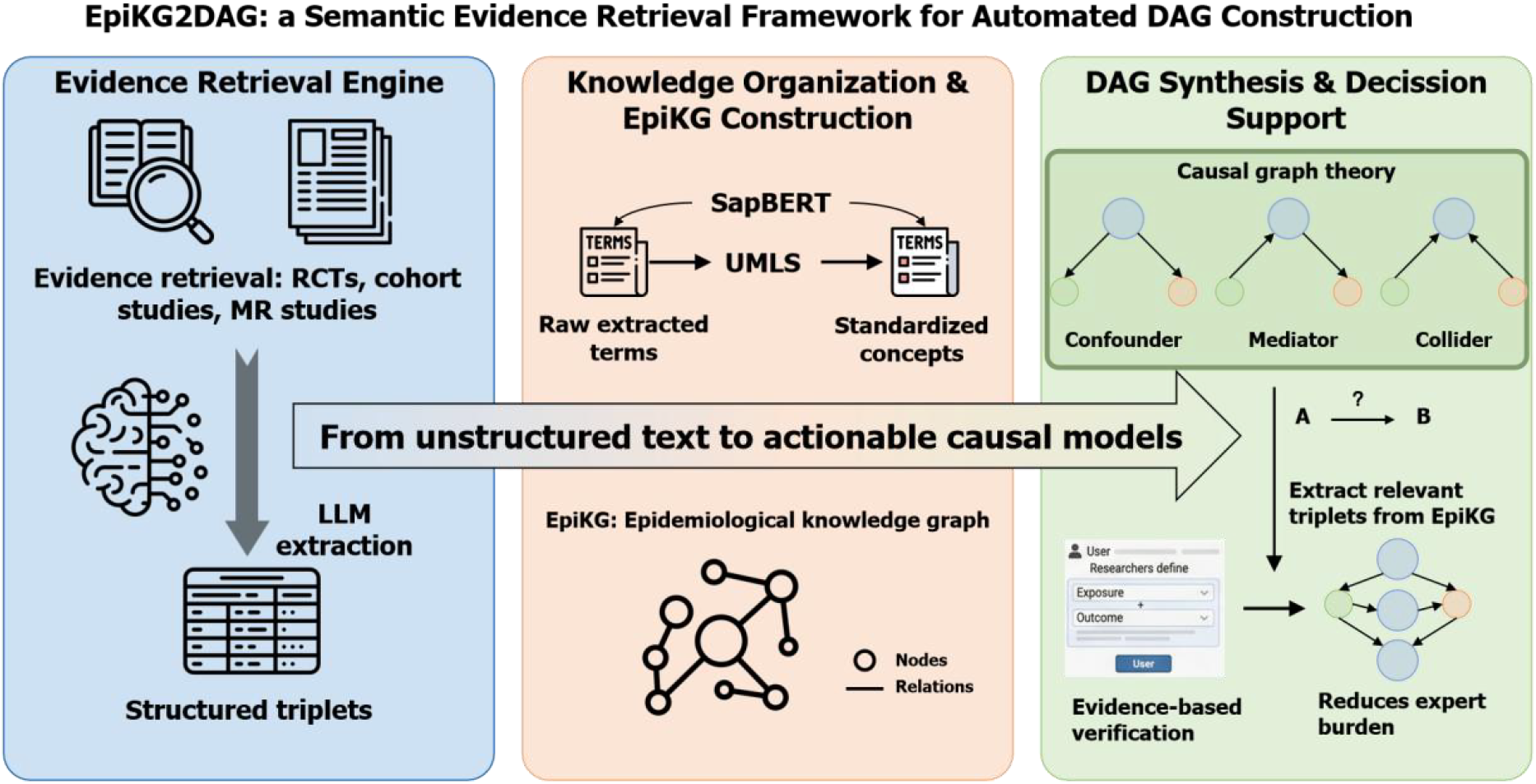
Technical Flowchart.

### Evidence Retrieval Engine

#### Data Source and literature Scanning

We conducted a search in the PubMed database. To establish a knowledge repository, we employed MeSH descriptors for “Cohort Studies,” “Randomized Controlled Trial,” and “Mendelian Randomization Analysis.” These study designs were selected because they are commonly used to generate or evaluate causal claims in epidemiology and therefore provide a relevant evidence base for candidate DAG construction. However, the inclusion of these designs does not imply that every extracted association represents definitive causal evidence; rather, study design information provides useful context for subsequent expert assessment. Furthermore, we used the MeSH Major Topic fields “C” (Diseases) and “F03” (Mental disorder) to ensure that the publications investigated at least one disease or symptom. Additionally, we applied the ISSN field to confine the publications to journals within the Nature Index (health science). After searching, a total of 74,992RCTs, 172,157 cohort studies and 1304 MR studies were obtained.

Subsequently, we excluded non-original research articles, such as reviews, editorials, and comments through the publication type field of the publication. Furthermore, given that the extraction process was based on abstracts, literature for which abstracts were not available was excluded from the study. Finally, a total of 70,189 RCTs, 118,294 cohort studies and 783 MR studies were included in the remaining dataset (Figure 2).

**Figure 2.**
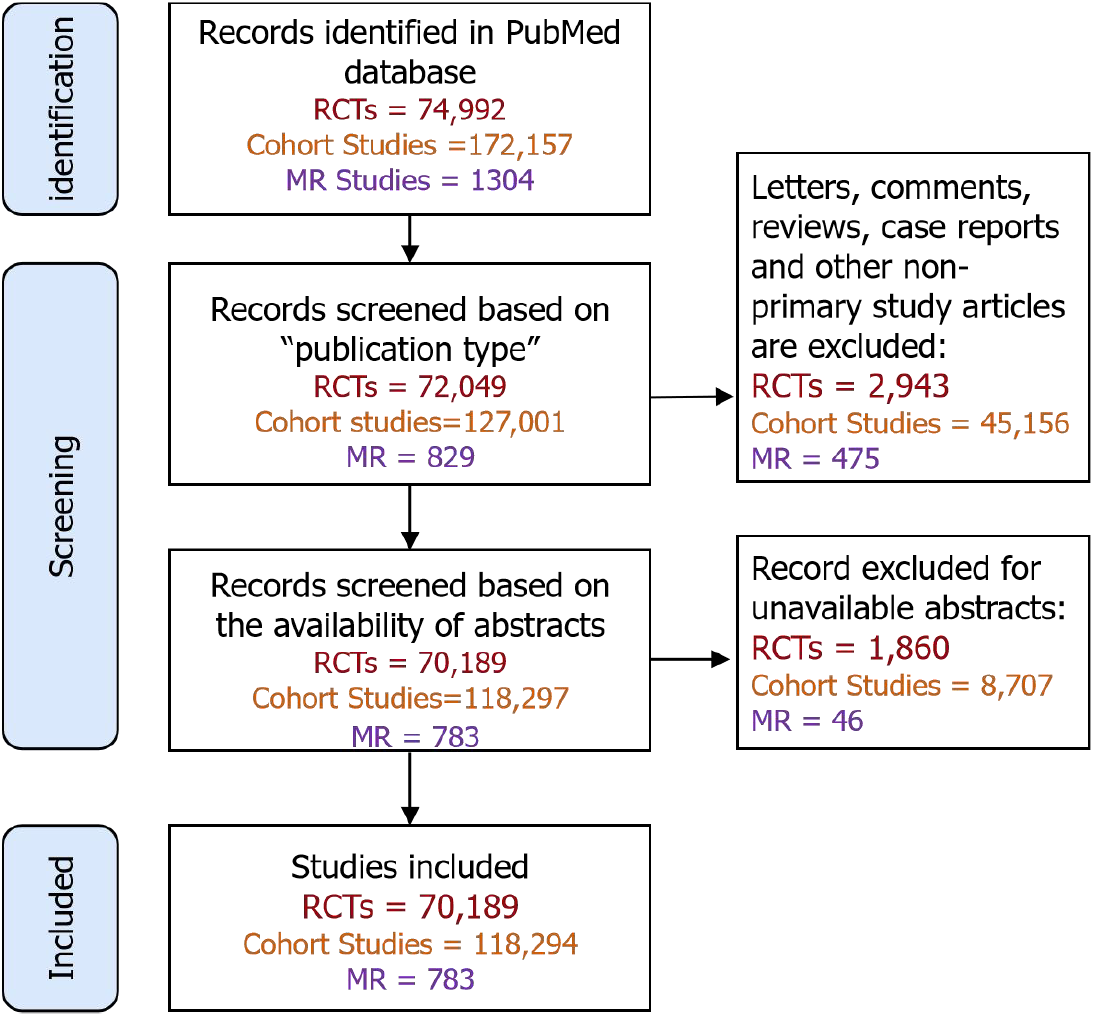
PRISMA flowchart for publication identification, screening and selection.

#### LLM-driven Information Extraction

We designed a prompt to extract the exposure, outcome, significance, direction, effect size, and values from the title and abstracts using LLM (Figure 3). Considering the study scope of causal inference in epidemiology, system prompt was set to “annotator in the domain of epidemiology.” A concise overview of each data element to be extracted was furnished. To facilitate entity standardization, it was specified that exposures and outcomes not be returned with abbreviations or acronyms. Direction was set to be returned as “increase,” “decrease” or “no change.” Significance was set to be returned as “positive” or “negative.” Value and 95% CI are only allowed to be returned as Arabic numerals. One-shot prompt was used to ensure that the LLM could understand the instructions and formats. The output format is set to JSON.

**Figure 3.**
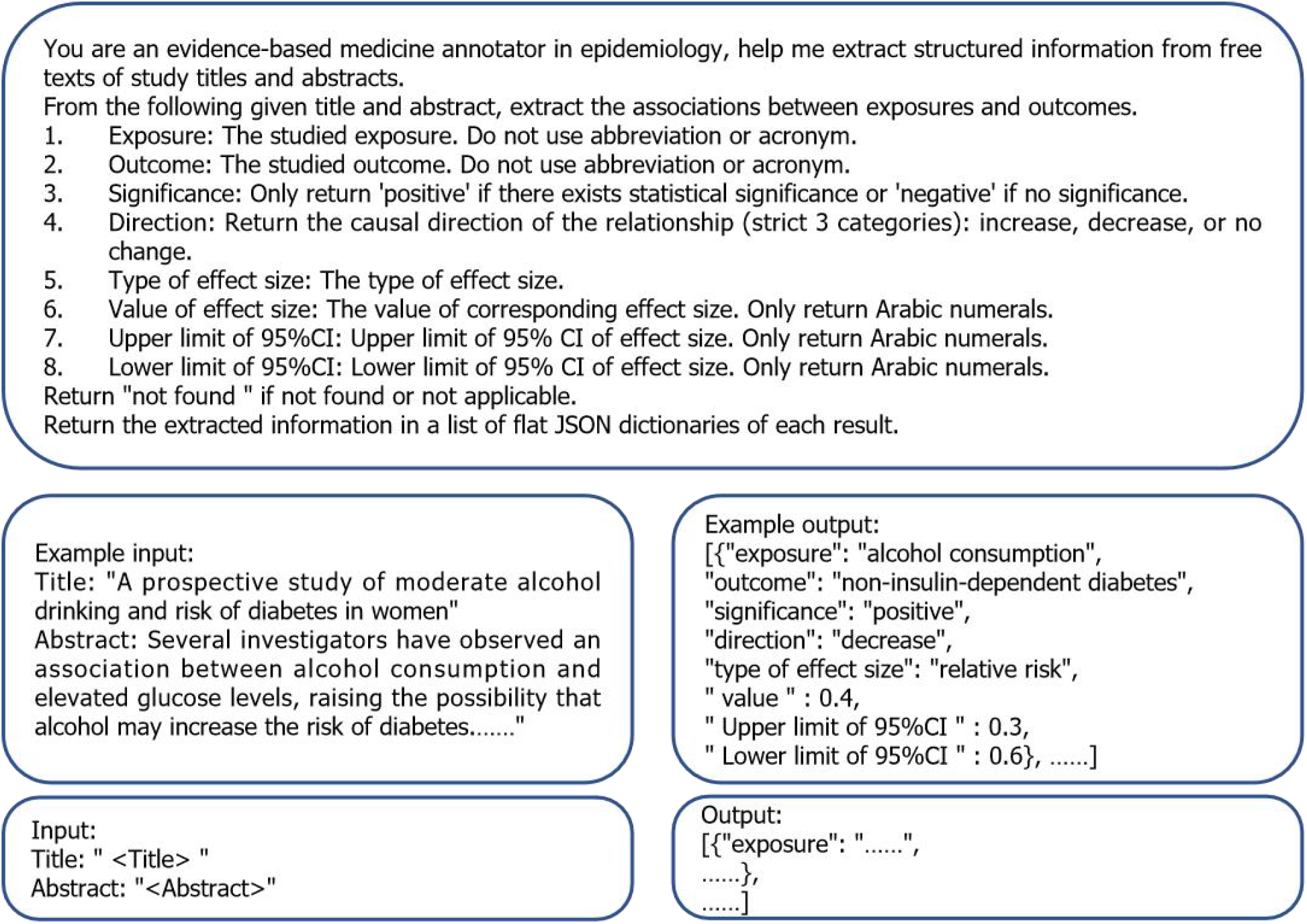
Prompt template.

**Figure 4.**
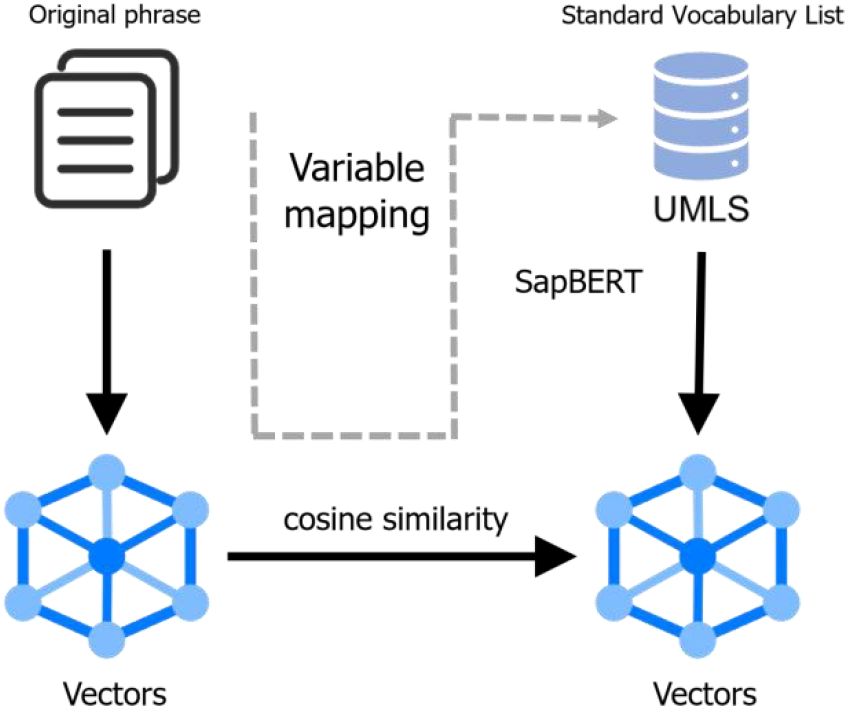
Semantic Normalization via SapBERT.

#### Pre-experiment and model selection

As a pre-experiment to evaluate extraction performance, both DeepSeek-V3(DeepSeek-AI et al., 2025) and ChatGLM-4-Plus(GLM et al., 2024) were utilized to extract information from the titles and abstracts of 150 randomly selected publications.

Performance was evaluated on a per-unit basis, with each extracted result classified as “correct,” “wrong,” or “supplement” (for findings missed by the LLM). To be marked “correct,” all six data columns—exposure, outcome, significance, direction, effect size type, and value—had to meet specific accuracy thresholds. While exposure and outcome were judged based on semantic consistency, significance, direction, and numerical values required exact matches. A more lenient criterion was adopted for the “type of effect size” (e.g., accepting “HR” for “adjusted HR”), as these variations did not substantively impact the causal interpretation.

Two reviewers independently evaluated the extracted results. For the results where two reviewers disagreed, the final decision was made by a senior reviewer. As shown in table 1, the precision, recall and F1-score were 0.914, 0.926, 0.920 (DeepSeek-V3) and 0.850, 0.934, 0.890 (ChatGLM-4-plus). Compared with ChatGLM-4-Plus, DeepSeek-V3 achieved a better balance between precision and recall, which is particularly important for downstream knowledge graph construction because false-positive associations may introduce spurious edges, whereas false-negative associations may reduce evidence coverage. Therefore, DeepSeek-V3 was selected as the extraction model for large-scale processing.

**Table 1.** Evaluation of two models. Precision, recall and F1-score were based on the final decision by the senior reviewer

| Model | Precision | Recall | F1 |
| --- | --- | --- | --- |
| Deepseek-chat-V3 | 0.914 | 0.926 | 0.920 |
| ChatGLM-4-plus | 0.850 | 0.934 | 0.890 |

### Semantic Normalization and EpiKG Construction

#### Entity Normalization with SapBERT

For entity standardization, we employed SapBERT(Abdulnazar et al., 2023), a PubMed Bert-based model specifically for entity linking tasks, to map the extracted exposures and outcomes to standardized UMLS concepts. SapBERT was selected because it is specifically optimized for biomedical entity linking and concept normalization, making it more suitable than general-purpose sentence embedding models for aligning heterogeneous clinical expressions. Compared with dictionary-based exact matching, embedding-based normalization can better handle lexical variation, synonyms, abbreviations, and paraphrased biomedical terms. Manual normalization, although potentially more accurate for small datasets, is not scalable for hundreds of thousands of extracted associations. Therefore, SapBERT provided a practical balance between scalability and biomedical semantic sensitivity. We used SapBERT to transform words into word vectors and compute cosine similarity to match. The cosine similarity threshold of 0.7 was used as a conservative heuristic to balance normalization coverage and semantic precision.

#### Knowledge Graph Architecture

The Epidemiological Knowledge Graph (EpiKG) was structured as a directed graph, with exposures designated as head nodes and outcomes as tail nodes. Edges were categorized based on the direction of the reported association as either “increasing” or “decreasing”. To prioritize statistically significant reported associations, associations originally classified as “no change” were excluded from the final graph, as they represent non-significant findings that do not contribute to actionable causal modeling. The weight of each edge represents the study frequency, reflecting the cumulative evidence for that specific relationship across the literature.

### DAG Synthesis and Decision Support

The framework utilizes causal graph theory to systematically identify structural motifs within the EpiKG, translating literature-derived associations into specific causal components. Candidate variables are categorized based on their topological relationship with a given exposure and outcome: confounders are defined as common causes that direct edges toward both variables; mediators are identified as intermediate nodes that receive an edge from the exposure and direct one toward the outcome; and colliders are identified as common effects receiving directed edges from both the exposure and the outcome. To ensure the structural integrity of the synthesized models, only edges representing statistically significant associations are utilized during motif identification, effectively filtering out non-significant findings that do not contribute to actionable causal reasoning. Crucially, as every edge within the EpiKG is anchored to its original publications, the framework facilitates seamless evidence tracing and verification. This architectural feature ensures the precision and completeness of the evidence base while significantly reducing the knowledge burden on experts by providing a verifiable, literature-anchored foundation for each identified relationship.

In the current implementation, EpiKG2DAG does not automatically force conflicting or bidirectional evidence into a single acyclic direction. Instead, direction-specific associations are preserved in EpiKG together with their source publications. When both *X* → *Y* and *X* → *Y* are detected, the relationship is flagged as bidirectional evidence for expert review before inclusion in the final candidate DAG.

## RESULTS

### LLM-driven Information Extraction

Following the systematic literature search and screening, the final dataset comprised 70,189 RCTs, 118,294 cohort studies, and 783 MR studies. These publications were processed using DeepSeek-V3 for automated information extraction, yielding an initial set of 620,217 associations. Among these, 141,361 associations were identified as “no change,” indicating a lack of statistical significance or discernible correlation. Given that the construction of EpiKG prioritizes statistically significant evidence to ensure the utility of downstream causal modeling, these non-significant findings were excluded. Consequently, a refined total of 478,856 significant associations were retained for inclusion in the knowledge graph. Each extracted association remains linked to its original PMID, establishing a robust foundation for subsequent evidence tracing and verification.

### Results of Semantic Normalization

Within the total pool of 478,856 significant associations, a unique set of 535,030 distinct terms (extracted from both exposure and outcome columns) was identified. Utilizing SapBERT for semantic alignment, 483,561 of these terms were successfully mapped to 145,295standardized UMLS concepts, achieving a robust normalization rate of 90.4%. This high degree of semantic consistency ensures that disparate terminologies across the literature are unified into a cohesive conceptual framework within the EpiKG.

### Statistics of EpiKG

Subsequently, a directed EpiKG was constructed, with exposures designated as the head nodes, outcomes as the tail nodes direction assigned as the type of edge, and the number of times exposure and ending were studied as the weight of edge. The EpiKG comprises a total of 145,295 nodes and 478,856 edges, of which 324,941 are of the “increasing” type, 153,915 of the “decreasing” type (Table 2).

**Table 2.** Statistics of the EpiKG.

|  | Number |
| --- | --- |
| **Publications** | 189,266 |
| **RCTs** | 70,189 |
| **Cohort studies** | 118,294 |
| **MR studies** | 783 |
| **Nodes** | 145,295 |
| **Edges** | 478,856 |
| **Increase** | 324,941 |
| **Decrease** | 153,915 |

In summary, an automated method was constructed for the extraction of exposure, outcome, significance, direction, type and value of effect size from the title and abstract based on a large language model. The method was found to be capable of extracting the information quickly and efficiently (P=0.914, R=0.926, F1-score=0.920) using DeepSeek, which greatly solves the problem of time- and energy-consuming manual reading.

### Case: DAG for the relationship between COVID-19 and acute kidney injury

We used the relationship between COVID-19-related exposure and acute kidney injury (AKI) as a case study because prior epidemiological studies have highlighted the importance of confounder adjustment when estimating the association between COVID-19 and AKI risk. Take acute kidney injury (AKI) as an example. After adjusting for confounders such as age, ethnicity, sex, hypertension, diabetes, heart failure, cardiovascular disease and obesity, the risk of AKI was greater in the unvaccinated group than in the vaccinated group, thus suggesting that the vaccine has a protective effect on patients. This study demonstrates the importance of adjusting for confounders in epidemiological studies.

Through EpiKG, we detected three node categories in the COVID-19-AKI relationship: (1) confounders (nodes with directed edges to both COVID-19 and AKI), (2) mediators (nodes receiving edges from COVID-19 and directing edges to AKI), and (3) colliders (nodes receiving directed edges from both COVID-19 and AKI). Through this approach, a candidate DAG can be obtained quickly (Figure 5). In this DAG, each edge links to the publication(s) from which the edge originated. This DAG can be used for rapid evidence examination and for further validation by human experts.

**Figure 5.**
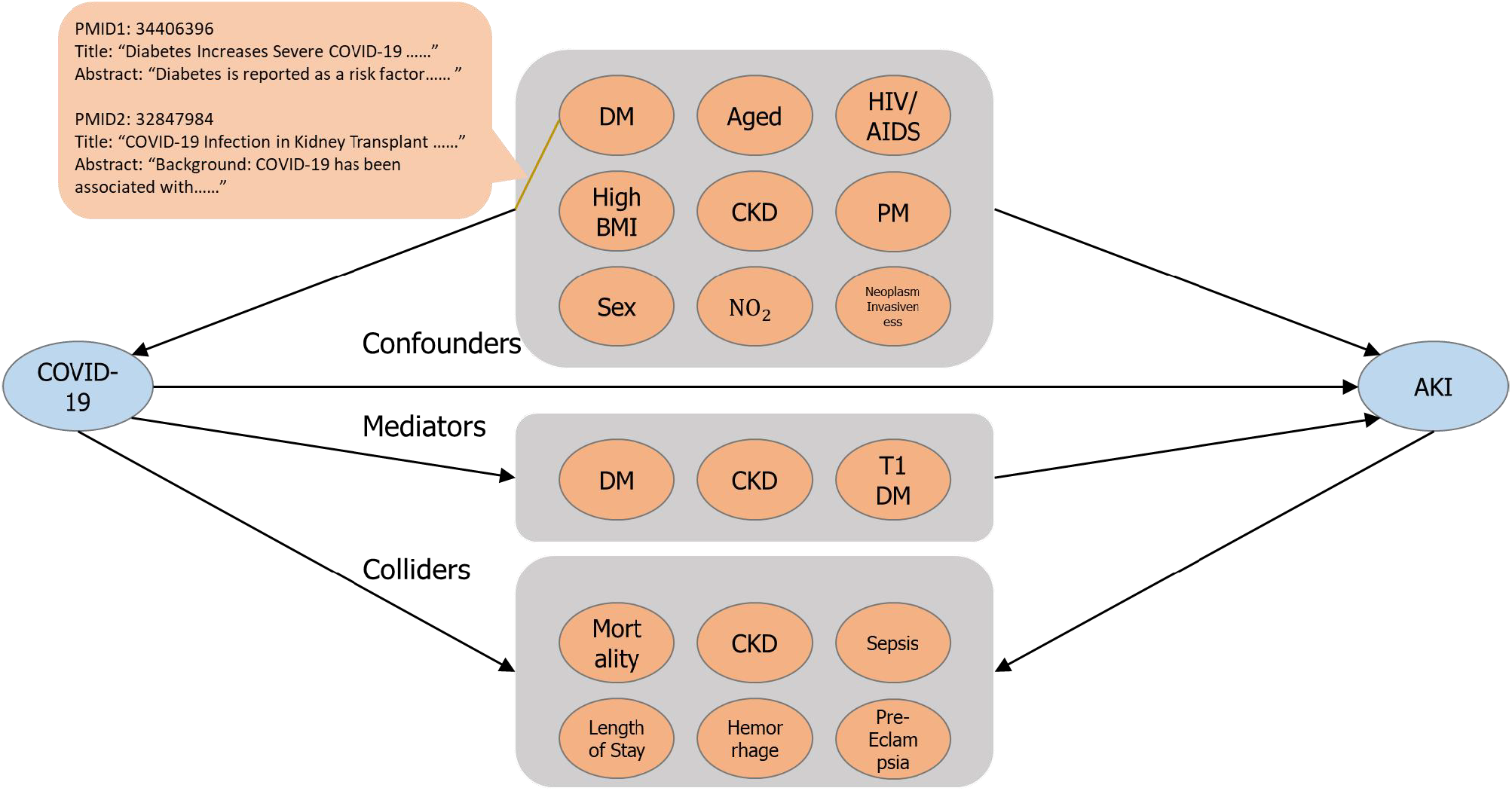
DAG for the relationship between COVID-19 and acute kidney injury. AKI: acute kidney disease. DM: diabetes mellitus. CKD: Chronic kidney disease. PM: particulate matter. T1DM: type 1 diabetes mellitus.

Among them, high BMI, sex and age are consistent with those mentioned in the literature(Pan et al., 2024). In addition, we also found unappreciated candidate confounders such as particulate matter, nitrogen dioxide, schizophrenia, neoplasm invasiveness, and so on. We retrieved the original publications through the evidence links. Take particulate matter and nitrogen dioxide as examples. One study suggested that exposure to particulate matter pollution and nitrogen dioxide was associated with a high risk of COVID-19(Kogevinas et al., 2021). Another study suggested that particulate matter pollution and nitrogen dioxide were risk factors for AKI(Lee et al., 2023). Taken together, these evidence-linked associations suggest that particulate matter and nitrogen dioxide may serve as candidate common causes of COVID-19 and AKI, and therefore warrant consideration as potential confounders in expert-refined DAGs. Similar evidence links were also found for neoplasm invasiveness (Kitchlu et al., 2019; Tian et al., 2020).

Moreover, in this DAG, we also quickly detected reverse causality, e.g., diabetes mellitus, diabetes mellitus type 1, chronic kidney disease, etc. Taking diabetes as an example, five publications have studied the development of diabetes mellitus after the diagnosis of COVID and found that the risk of developing diabetes is significantly higher for a period of time following the diagnosis of COVID-19(Ayoubkhani et al., 2021; Rezel-Potts et al., 2022; Wong et al., 2024; Xie & Al-Aly, 2022; Xiong et al., 2023). In turn, three more studies have shown that diabetes is significantly associated with a higher risk of developing COVID-19(Diedisheim et al., 2021; Elias et al., 2020). Therefore, from the perspective of correlation studies, it is difficult to explain which is the cause and which is the effect, and human expert validation is still needed.

## DISCUSSION

### Mitigating the “Evidence Retrieval Gap” in Causal Modeling

The primary contribution of this study is addressing the critical evidence retrieval gap in observational research. While systematic literature synthesis is foundational to evidence-based medicine, our research highlights that manual DAG construction is frequently plagued by an opaque and non-systematic knowledge acquisition process. By introducing EpiKG, we demonstrate that framing DAG development as an automated knowledge integration task can significantly mitigate the subjectivity and inefficiency inherent in traditional human-centric approaches. The inclusion of diverse study designs—comprising RCTs, Cohort Studies, and MR—ensures a multi-dimensional evidence base that minimizes retrieval bias and captures a broader spectrum of causal candidates.

### Comparison with Existing Approaches

EpiKG2DAG can be better understood by comparing it with two related approaches: ESC-DAGs(Ferguson et al., 2019) and SemMedDB-based methods(Bai et al., 2024). ESC-DAGs synthesizes variables considered in previous studies of a target exposure–outcome relationship into a shared DAG, which improves the transparency of evidence use. However, it is still largely bounded by variables and DAG structures already reported in existing target studies; given the limited transparency and inconsistency of DAG construction in medical research, this may inherit omissions from prior studies. By contrast, EpiKG2DAG retrieves and organizes a broader literature-derived association network related to both the exposure and outcome, enabling candidate common causes, intermediate variables, and common effects to be identified by graph topology.

EpiKG2DAG also differs from SemMedDB-based approaches. SemMedDB provides PubMed-scale UMLS-normalized subject–predicate–object semantic predications for biomedical knowledge discovery, but its broad-coverage triples may include redundant, generic, or semantically broad concepts that are not optimized for epidemiological DAG construction. In contrast, EpiKG2DAG extracts exposure–outcome associations from selected empirical study designs and retains direction, statistical significance, effect-size information, and PMID-level evidence links, making EpiKG more directly suited to evidence-anchored candidate DAG generation.

### Discovery of Unappreciated Confounders

The practical utility of the EpiKG framework extends beyond the replication of established medical knowledge to the discovery of unappreciated or non-obvious causal factors. In our case study on the relationship between COVID-19 and Acute Kidney Injury (AKI), the framework successfully identified established confounders such as age, diabetes, and obesity. More importantly, it detected novel candidates, such as air pollution (particulate matter and nitrogen dioxide) and neoplasm invasiveness, which were supported by empirical literature but often omitted from expert-driven models. By scanning the entire Nature Index corpus, EpiKG provides a more comprehensive and systematic view of the causal landscape than is possible through manual review.

### Reducing Knowledge Burden through Evidence Traceability

EpiKG serves as a transparent decision-support tool that reduces the knowledge burden of manual DAG construction by automating the extraction, normalization, and organization of large-scale epidemiological associations. Unlike traditional workflows that require labor-intensive literature screening, EpiKG links each identified edge to its source publications, allowing users to rapidly trace and verify the evidence behind candidate relationships. In practice, EpiKG2DAG supports an expert-in-the-loop workflow: researchers can inspect evidence-linked edges for a specified exposure–outcome pair, evaluate study relevance, temporal ordering, possible reverse causality, and consistency across publications, and then decide whether candidate variables should be retained, removed, or reclassified before formal causal analysis.

### Limitations and Future Directions

Despite its performance, several limitations remain. First, the current extraction relies on titles and abstracts, which may omit important information reported only in the full text, including subgroup-specific findings and complete numerical effect estimates. Such abstract-level incompleteness may affect downstream DAG validity by introducing missing edges. Therefore, the candidate DAGs generated by EpiKG2DAG should be interpreted as evidence-anchored starting points for expert review rather than final causal models.

Future work will incorporate full-text corpora and section-aware extraction, particularly from the paragraph, figures and tables in the Results sections, to capture subgroup analyses and more complete effect estimates. We will also compare abstract-only and full-text-based outputs to evaluate how additional contextual evidence changes the structure and validity of generated candidate DAGs.

## CONCLUSION

In conclusion, EpiKG2DAG establishes a systematic framework for generating evidence-anchored candidate DAGs from biomedical text, addressing the evidence retrieval gap in observational research. By integrating LLM-driven association extraction, SapBERT-based semantic normalization, and graph-theoretic motif identification, the framework transforms 189,266 abstracts into a large-scale Epidemiological Knowledge Graph for candidate causal structure generation. The COVID-19–AKI case study shows that EpiKG2DAG can replicate established confounders and identify less obvious candidates, such as air pollution, while preserving PMID-level evidence traceability. Overall, EpiKG2DAG provides a scalable decision-support tool for evidence-informed DAG development and expert-guided causal modeling.

## GENERATIVE AI USE

We confirm that we did not use generative AI tools/services to author this submission.

## Data Availability

All data produced in the present study are available upon reasonable request to the authors

## ACKNOWLEDGMENTS

This study is supported by the National Key R&D Program for Young Scientists (Project number 2022YFF0712000 to JD)

